# A Systematic Review of Sex Differences in Postoperative Nausea and Vomiting

**DOI:** 10.64898/2026.06.23.26356213

**Authors:** Carol Huang, Nethuli Kolugala, Jasmine Druskovich, Mikaela Law, Cameron Wells, Chris Varghese, Michelle R Wise, Greg O’Grady

## Abstract

**Background:** Postoperative nausea and vomiting (PONV) is a common consequence of anaesthesia, affecting up to 30% of postoperative patients. Female sex is one of the strongest risk factors for PONV, yet no dedicated analysis has examined how this association varies across surgical settings and timepoints. This systematic review and meta-analysis aimed to quantify sex differences in PONV incidence across different surgical contexts.

**Methods:** A systematic search was conducted using PRISMA guidelines across Medline and Embase from inception to September 1, 2025. Eligible studies were observational cohort studies (n≥500) of adult patients that conducted multivariate regression analyses including sex as a variable. Two reviewers independently screened, extracted data, and assessed risk of bias using ROBINS-E. A random-effects meta-analysis was performed. Subgroup analyses and multiple sensitivity analyses were completed.

**Results:** From 4620 identified studies, 23 met the inclusion criteria, including 462,828 patients across various surgical settings and specialties (52% female). The pooled incidence of PONV was 21% (95% CI[16-27%]), with high heterogeneity (I^2^=99.9%). Meta-analysis confirmed females had a higher risk of developing PONV compared to males (pooled OR=2.40, 95% CI[2.06-2.79], I^2^=93.1%, p<0.0001). Sensitivity analyses confirmed robustness of pooled estimates. Subgroup analyses demonstrated consistently elevated risk of PONV for females at all three timepoints (PACU, 24-hour, 48-hour post-operative) and across studies including and excluding female-only surgeries. Studies were generally at high risk of bias.

**Discussion:** Female sex is a strong risk factor for PONV across surgical settings. Further research into precise subgroups, underlying mechanisms of sex-differences, and the use of prophylaxis may help improve this inequity.

## Introduction

Postoperative nausea and vomiting (PONV) is a common consequence of anaesthesia, affecting up to 30% of the general population and up to 80% of high-risk patients (1). PONV impacts patient recovery, experience, and healthcare resource utilisation, and represents a significant financial and logistical burden in perioperative care (2). Multiple factors contribute to PONV, encompassing patient, procedural, and anaesthetic characteristics.

A 2012 systematic review by Apfel *et al*. analysed risk factors of PONV and found female sex to be the strongest patient-specific predictor (3), with gynaecological and breast procedures associated with high risk, potentially indicating confounding by sex (4). However, while female sex is a risk factor, there have been no previously focused meta-analyses of this association, including how the risk varies across surgical contexts and timepoints. In addition, while contemporary guidelines emphasise the importance of PONV prophylaxis (1), nearly 95% of female patients are under-treated with PONV prophylaxis (5). Nausea and vomiting experienced by females is therefore an under-addressed area of perioperative care, requiring an improved understanding of why sex differences exist both in the development of PONV and its management.

This study therefore aimed to systematically review the perioperative literature to quantify sex differences in the incidence of PONV and define potential contributing factors.

## Methods

This systematic review was conducted and reported in accordance with the Preferred Reporting Items for Systematic Reviews and Meta-Analyses (PRISMA) statement (6). The review protocol was developed apriori and prospectively registered (CRD420261332547).

### Search Strategy

A systematic search was conducted on Medline (OVID) and EMBASE from the beginning of the review to September 1, 2025. The following terms were used for the search: “Regression Analysis OR Multivariate Analysis OR Discriminant Analysis” AND “Female OR Male” AND “Sex factors OR Risk factors” AND “Postoperative Nausea and Vomiting”. Details of the full search strategy can be found in Appendix A.

For full-text articles that could not be retrieved, the corresponding author of the article was contacted to request access. Relevant reference lists of accepted articles were manually searched to identify any additional relevant articles that were not captured in the database search. All articles found were extracted to a systematic review software (Rayyan), where all duplicates were removed.

### Eligibility Criteria

Prospective and retrospective cohort studies examining PONV as a primary or secondary outcome with multivariable regression analyses examining the impact of sex on PONV occurrence were included. Studies that measured PONV beyond 48 hours were excluded, to ensure that outcomes reflected true PONV rather than other causes of nausea and vomiting. Only full-text articles were included; case reports, expert opinions, and editorials were excluded. Due to language restrictions and potential inconsistencies with translations, eligible studies were all published in English. All studies published before 1960 were excluded, when older inhalational anaesthetic agents with known increased risk of PONV were still widely used (7).

Included studies were required to have a minimum sample size of 500 participants. Randomised controlled trials were excluded, as the protocolised interventions and selected populations in such trials may not accurately reflect PONV incidence in routine clinical practice. Studies were restricted to those studying adult participants aged 18 years or older, with all studies that included paediatric participants excluded unless adult-only data could be extracted. All studies involving animal subjects were excluded. Eligible studies must have included both male and female participants to enable comparisons.

For consistency across comparisons, only studies including patients receiving operations under general anaesthesia (GA) were included, with studies of sedation, spinal anaesthesia, or epidural-only modes of anaesthesia excluded. All surgical settings and specialities were eligible, including inpatient, outpatient, emergency, elective, and surgeries of all specialities, except dental procedures due to their distinct perioperative context.

To ensure meaningful interpretation of effect estimates, only studies in which PONV was a primary or secondary outcome were included, reducing the risk of extracting sex coefficients from models not designed to assess this association. Additionally, only studies employing multivariable regression analyses with covariate adjustment that included sex (biological male or biological female) were included.

### Protocol Deviation

In the original protocol, only prospective cohort studies were eligible for inclusion. However, during the study selection process, this criterion was expanded to include retrospective cohort studies with a minimum sample size of 500 participants. This deviation was made after preliminary screening identified a number of large, methodologically robust retrospective studies, including retrospective analyses of prospectively collected data. To maintain rigour, retrospective studies were only included if they met all other inclusion criteria (as defined above), and the same sample size threshold (n ≥ 500) was applied to minimise sampling bias and enhance reliability. Additionally, the protocol specified that included studies must explicitly define participants as biological female or biological male. In practice, this was not consistently reported. Therefore, studies were included if they reported both female and male participants.

### Screening and Study Selection

Two independent reviewers (CH and NK) screened all studies. First, each reviewer independently screened the titles and abstracts of all identified studies for the eligibility criteria. Any articles whose relevance could not be determined by a title and abstract screening alone were included for further review. Second, a full-text review of the remaining articles was performed by CH and NK. Results were discussed to achieve consensus, with a third independent reviewer utilised to resolve disagreements if they arose.

### Variables

Data were extracted from the final included articles by two independent reviewers (CH and NK). All extracted data were stored using a standardised charting form that was piloted by the two reviewers prior to data extraction. The following variables were extracted: publication details, study design, details on the population and sample (sample size, sex distribution, age of participants, and location of the study), type of surgery, type of anaesthetics, reported outcomes (PONV as the primary interest), and key findings (statistical outcomes). The primary data of interest extracted from each included study were the odds ratio (OR) and its associated 95% confidence interval (CI) for the association between female sex and the risk of developing PONV, as reported in multivariable regression analyses.

Reported OR where females were used as reference were mathematically converted, by both independent reviewers, to ensure that males were consistently the reference sex.

### Risk of Bias Assessment

Risk of bias was assessed using ROBINS-E, and was performed by two independent reviewers (CH and NK) (8). This tool evaluates seven domains of bias and provides an overall judgement regarding risk of bias, the likely direction of bias, and potential impact on the validity of the study’s conclusions (8).

### Statistical Analysis

The meta-analysis was performed in R (version 2025.05.1+513) using the *meta* package. A random-effects model was used to account for the high level of between-study heterogeneity. The adjusted odds ratios extracted from the studies’ multivariate regression analyses, along with their associated 95% CIs, were pooled in the model. Heterogeneity was quantified using the I² statistic, which represents the percentage of total variation across studies attributable to between-study differences rather than chance. Pooled study-level incidences were calculated for studies that reported multiple overall PONV incidences for different subgroups. Two studies reported more than one dataset (9,10). Due to this, analyses were performed on 25 unique datasets rather than 23.

Sensitivity analyses, including a Baujat plot and leave-one-out sensitivity analysis using a random effects model (*metainf*), were conducted to evaluate the level of influence the individual studies had on the pooled effect to better understand the level of heterogeneity.

### Subgroup Analyses

Subgroup analyses stratified by the timing of PONV measurement and the type of surgery included were performed. The first subgroup analysis stratified the results by the time of PONV reported: during stay in the post anaesthesia care unit (PACU), 24 hours post-operatively, and 48 hours post-operatively. The second subgroup analysis assessed whether the inclusion of surgical procedures performed predominantly or exclusively on females, such as obstetric, gynaecological, or breast cancer surgeries, accounted for the observed sex difference in PONV. Because these procedures may independently carry higher PONV risk, their inclusion could inflate the pooled estimate for females. Studies were categorised as “includes female-specific surgeries”, “excludes female-specific surgeries”, or “unclear” where surgical details were insufficiently reported. Studies in the “includes female-specific surgeries” subgroup still included both male and female participants, as female-specific procedures were analysed alongside non-sex-specific surgeries.

## Results

### Study Selection

Database and hand searching revealed 4,620 results, of which 23 studies met the inclusion criteria for analysis. Study screening with reasons for exclusions are detailed in the PRISMA diagram (Figure 1).

**Figure 1.**
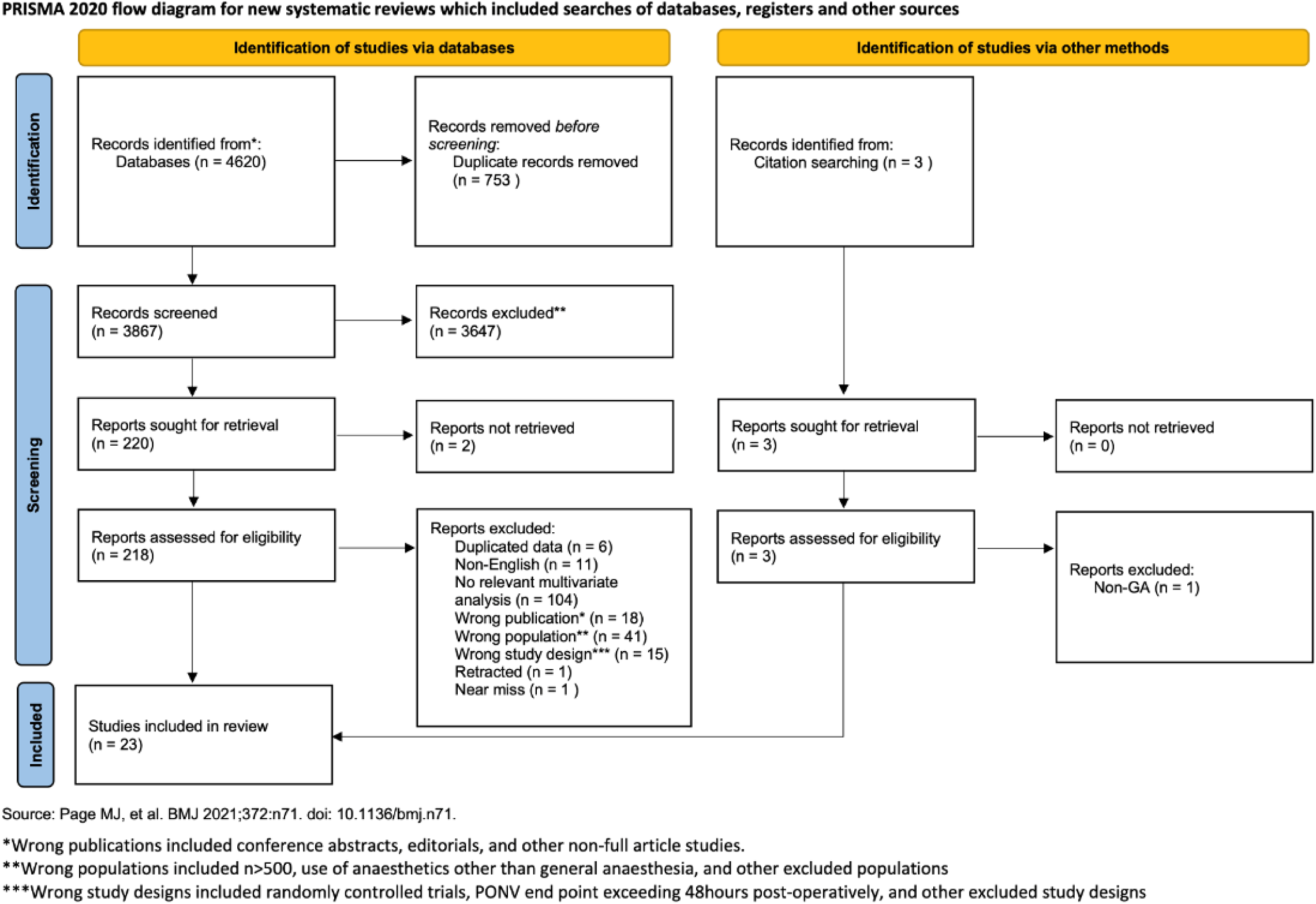
Preferred Reporting Items for Systematic Reviews and Meta-Analyses (PRISMA) 2020 flow diagram for study screening and inclusion.

Full study characteristics of the included studies are presented in Table 1. The majority of included studies had a retrospective design (n=12, 52.2%), with the remainder including nine prospective studies (39.1%) and two retrospective reviews of prospective data (8.7%). The included studies were predominantly conducted in East Asia (n=11, 47.8%), followed by Europe (n=5, 21.7%) and the United States (n=4, 17.4%). One study was conducted in Australia and one study in South Africa. More than half of the included studies (n=15, 65.2%) were published within the past 10 years (since 2016). Most studies included a wide range of different surgical specialities. The studies were heterogeneous in nature, both in terms of the populations included and the measurement of PONV. PONV outcomes were assessed at 24 hours (n=13) and 48 hours (n=6) post-operation, as well as during recovery in the PACU (n=4).

**Table 1.** Characteristics of included studies. GA = general anaesthesia; GA + combined = general anaesthesia and combined anaesthetic techniques; PACU = post-anaesthesia care unit; Various = wide range of surgical specialties. PONV end points were assessed in PACU, at 24 hours postoperatively, or at 48 hours postoperatively.

| Author (Year) | Country | Study Design | Sample Size | Proportion of Females (%) | Type of Anaesthetic | Type of Surgery | End Point of PONV |
| --- | --- | --- | --- | --- | --- | --- | --- |
| Apfel, C.C (1999) | Finland + Germany | Prospective Cohort Study | 2722 | 48.2 | GA | Various | 24hr |
| Apfel, C.C (2012) | USA | Prospective Cohort Study | 1913 | 65.9 | GA | Various | 48hr |
| Brettner, F (2016) | Germany | Register Based Cohort Study | 2617 | 47.3 | GA + combined | Various | PACU |
| Chae, D (2020) | South Korea | Retrospective Study of Prospective Data | 22144 | 46 | GA | Various | 48hr |
| Choi J.B. (2014) | South Korea | Retrospective Cohort Study | 1878 | 51 | GA | Various | 48hr |
| Douville, N.J (2025) | USA | Prospective Cohort Study | 64,523 | 51.8 | GA + combined | Various | PACU |
| Eberhart, L H (2000) | Germany | Prospective Cohort Study | 1444 | 56.7 | GA | Various | 24hr |
| Huang, C (2025) | China | Retrospective Cohort Study | 5606 | 57.8 | GA + combined | Elective Thoracoscopic Assisted Lung Resection | 24hr |
| Kao, S.C (2017) | Taiwan | Retrospective Cohort Study | 4143 | 58.4 | GA | Major Abdominal | 48hr |
| Kim J.H (2020) | South Korea | Retrospective Cohort Study | 103561 | 60.75 | GA | N/A | 24hr |
| Kratt, K.M (2019) | USA | Retrospective Cohort Study | 794 | N/A | GA | Ophthalmology (Strabismus) | PACU |
| Kwon Y.S (2020) | South Korea | Retrospective Cohort Study | 202439 | 62.4 | GA | N/A | 24hr |
| Li, N (2021) | China | Observational Cohort Study | 3223 | 37.9 | GA | Various | 24hr |
| Mc Loughlin, S (2019) | Argentina | Retrospective Observational Study | 753 | 52 | GA | Elective Colorectal | 48hr |
| Myles, P.S (1996) | Australia | Retrospective Observational Study | 4173 | 45 | GA | Various | 24hr |
| Nakagawa, M (2008) | Japan | Prospective Cohort Study | 1070 | 52.2 | GA | Elective Cancer | 24hr |
| Rodseth, R.N (2010) | South Africa | Prospective Observational Study | 800 | 50 | GA | Various | 24hr |
| Stamer, U.M (2019) | Germany + Switzerland | Prospective Observational Study in Two Independent Cohorts | A = 918<br>B = 1663 | A = 33.7<br>B = 58.3 | GA | A = Mainly Urological + Major Abdominal<br>B = Various | 24hr |
| Suhre, W (2020) | USA | Retrospective Observational Study | 27388 | 55.7 | GA | Various | PACU |
| Suzuki, Y (2023) | Japan | Retrospective Observational Study | 666 | 65.17 | GA | Various | 24hr |
| Wallenborn, J (2007) | Germany | Prospective Observational Study | 625 | 43.04 | GA | Lumbar Disc | 24hr |
| Wu, Y.H (2015) | Taiwan | Retrospective Observational Study | 992 | 58.4 | GA | Various | 24hr |
| Yi, M.S (2016) | South Korea | Retrospective Study of Prospective Data | 6773 | 60.1 | GA | Various | 48hr |

The final 23 studies comprised 462,828 patients, of whom 52% were female. The included studies were published between 1996 and 2025. The sample size ranged from 625 to 202,439 patients, with a pooled incidence of PONV of 21% (95% CI [16-27%]). There was a very high level of heterogeneity (I^2^=99.9%), as shown in Figure B-1 (Appendix B). One study did not report the total incidence of PONV.

### Primary Meta-Analysis

In all studies except one by Kratt *et al*. (11), females had a higher risk of developing PONV compared to males. A pooled analysis demonstrated that females had a significantly higher risk of developing PONV compared to males (pooled OR=2.40, 95% CI[2.06 - 2.79], I^2^=93.1%, p<0.0001, Figure 2).

**Figure 2.**
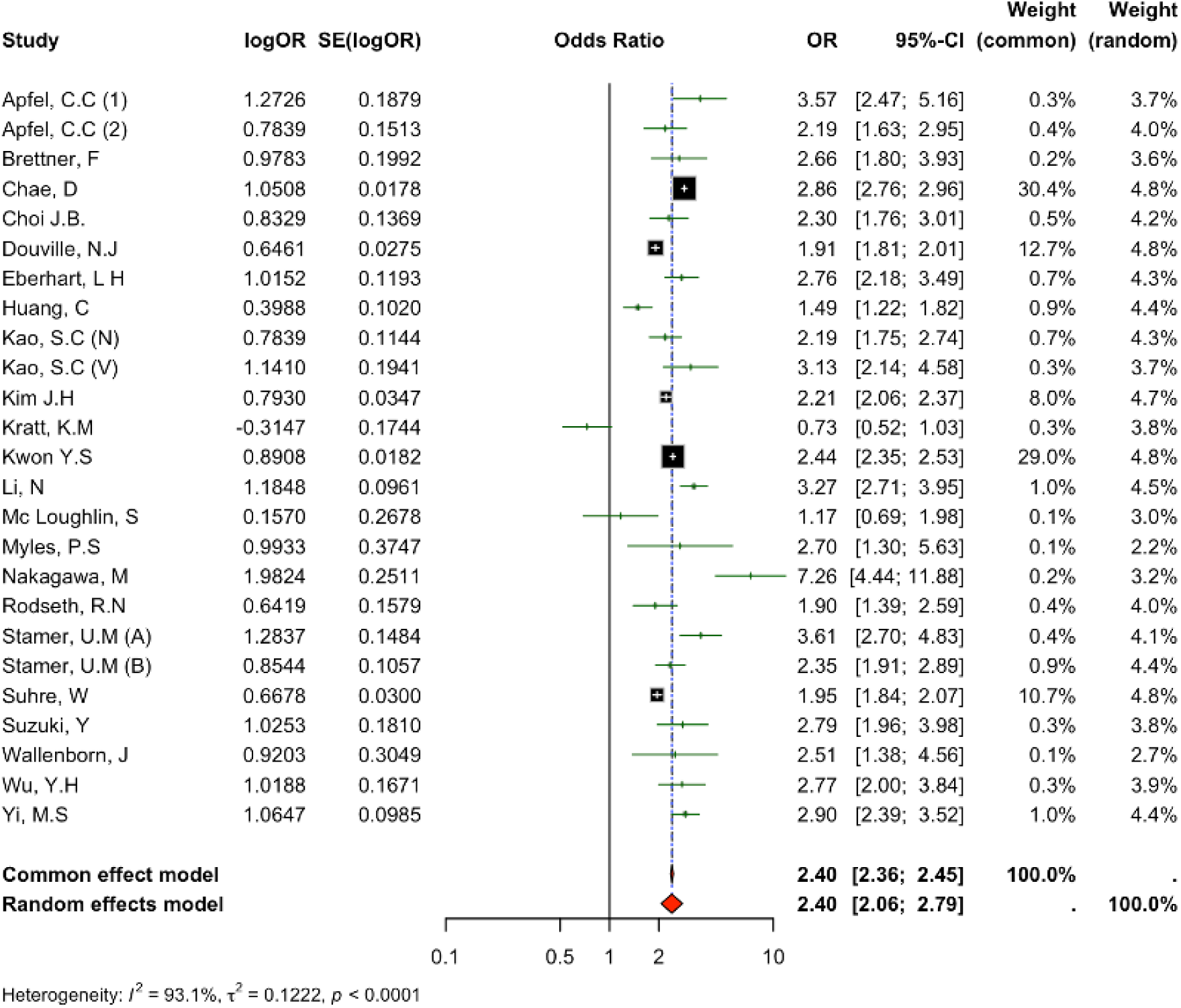
Meta-analysis of the association between female sex and PONV. Forest plot showing pooled adjusted OR and 95% CI using common-effect and random-effects models. Males were the reference sex; ORs >1 indicate increased PONV risk in females. Two studies by Apfel et al. are labelled (1) and (2). Kao et al. reported separate nausea and vomiting datasets, labelled (N) and (V), respectively. Stamer et al. included two independent cohorts, labelled (A) and (B).

### Subgroup Analyses

#### Timing of PONV

When studies were stratified by timing of PONV measurement, females remained at elevated risk for developing PONV at all three time points: in PACU OR=1.65 (0.98-2.78) (n=4), at 24-hour OR=2.70 (2.36-3.22) (datasets n=14), and 48-hour end point measures OR=2.44 (2.07-2.88) (n=7) (Figure 3). When PONV was measured in PACU, this resulted in a lower pooled OR compared to both the 24-hour and 48-hour endpoint measures. Interestingly, the PACU subgroup included the only study, Kratt *et al.*, which reported a lower incidence of PONV for females than males (11). Heterogeneity remained high in all three subgroups at I^2^ > 70%.

**Figure 3.**
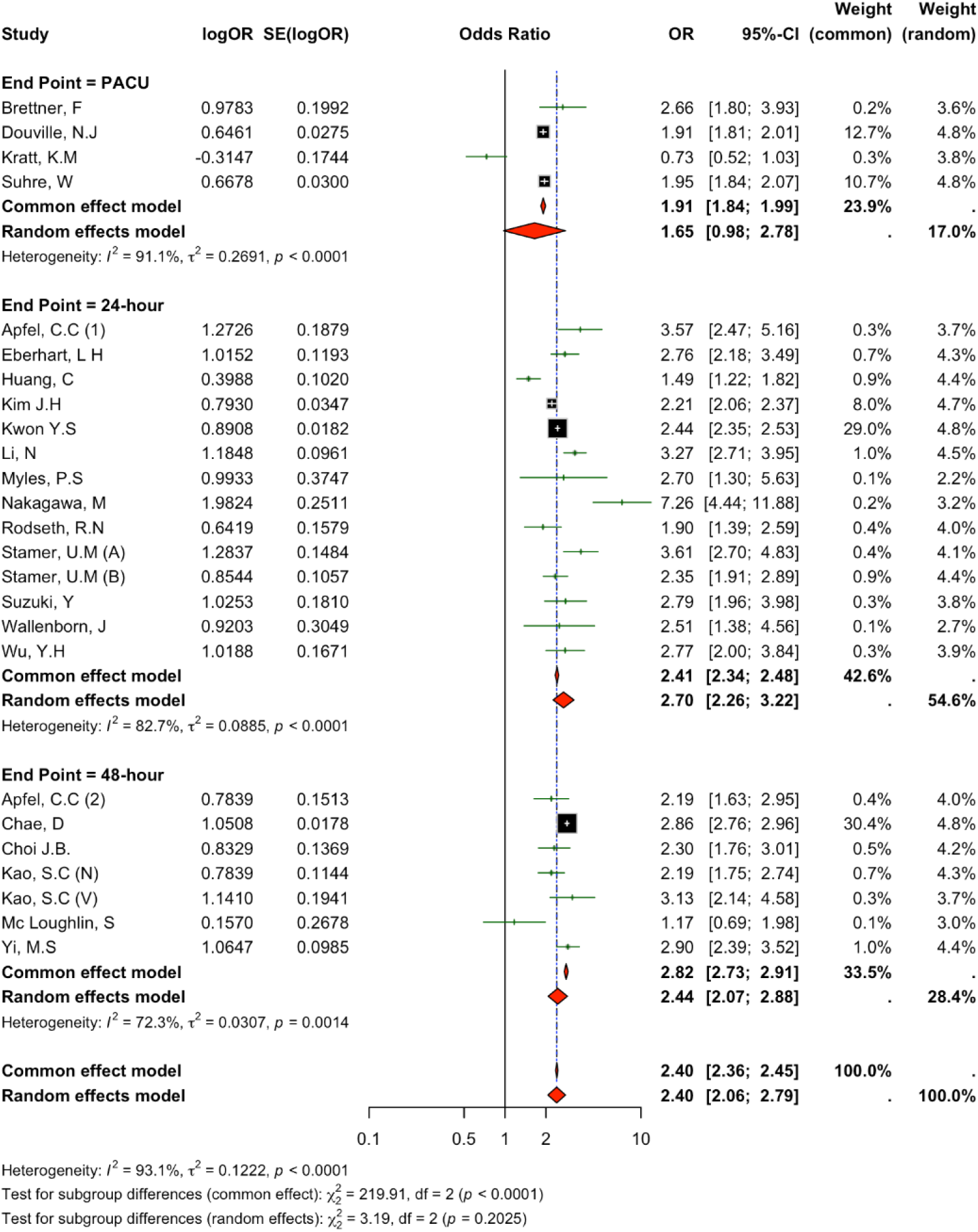
Subgroup analysis by end point of PONV measurement. PACU (n=4), 24 hours postoperatively (n=14), and 48 hours postoperatively (n=7). Two studies by Apfel et al. are labelled (1) and (2). Kao et al. reported separate nausea and vomiting datasets, labelled (N) and (V), respectively. Stamer et al. included two independent cohorts, labelled (A) and (B).

#### Female Exclusive Surgeries

As per the methods, to assess whether the observed sex difference was driven by the inclusion of surgeries performed predominantly on females, studies were stratified by whether they included female-specific procedures (e.g., gynaecological, obstetric, or breast surgeries). The pooled OR remained elevated in all three subgroups: studies that included female-specific surgeries alongside non-sex-specific surgeries (OR=2.70; 95% CI 2.22–3.29; n=9), studies that excluded female-specific surgeries (OR=2.07; 95% CI 1.44–2.97; n=9), and studies with unclear surgical classification (OR=2.35; 95% CI 2.06–2.68; n=7; Figure 4). The persistence of elevated risk in studies excluding female-specific procedures indicates that the sex disparity in PONV is not solely attributable to surgical type. Heterogeneity remained high across all three subgroups (I²>80%). Of note, Myles et al. reported *”no obstetric service and very little gynaecological surgery”* (12) and was therefore classified under excludes female-specific surgeries; Stamer et al. included two independent cohorts, of which Cohort B included gynaecological and breast surgeries while Cohort A did not include any female-exclusive surgeries (9).

**Figure 4.**
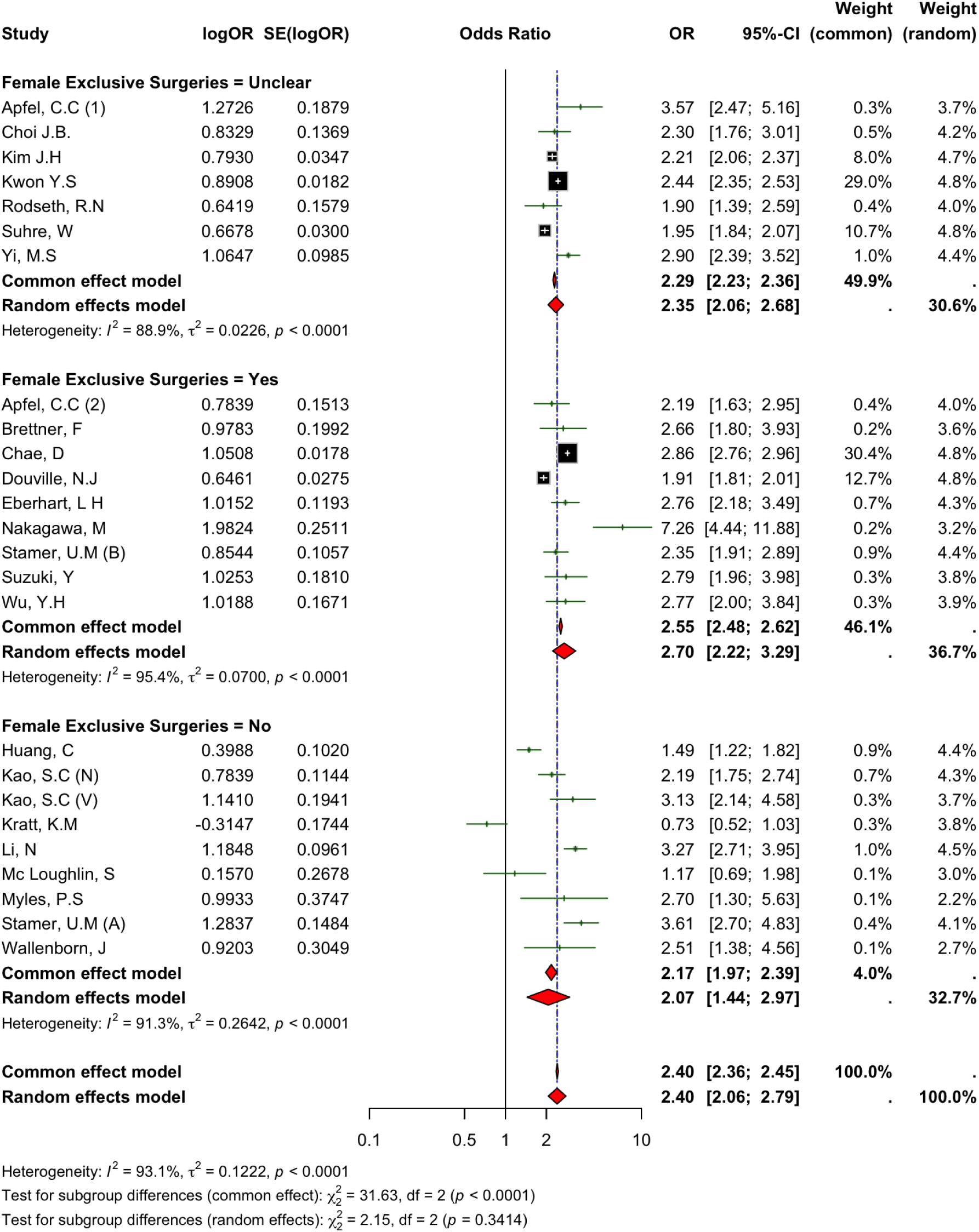
Subgroup analysis by inclusion of female-only surgeries. Includes female-only surgeries (n=9), excludes female-only surgeries (n=9), and unclear (n=7). Two studies by Apfel et al. are labelled (1) and (2). Kao et al. reported separate nausea and vomiting datasets, labelled (N) and (V), respectively. Stamer et al. included two independent cohorts, labelled (A) and (B).

### Use of Prophylactic Antiemetics

Reporting of prophylactic antiemetic use was inconsistent across included studies and was frequently insufficient to allow meaningful comparison between sexes. While several studies described the use of common agents such as dexamethasone and 5-HT3 receptor antagonists, most did not stratify administration by sex, limiting the ability to determine whether females received different levels of prophylaxis compared to males. In a small number of studies, prophylaxis was standardised or universally administered, whereas others reported variable or clinician-dependent use, further contributing to heterogeneity. Only limited data were available on sex-specific effectiveness of antiemetic prophylaxis, with one study by Brettner *et al*. suggesting no significant modification of PONV risk in females despite prophylactic treatment (13). Overall, the lack of consistent, sex-disaggregated reporting of prophylaxis use and efficacy precluded robust analysis of whether differences in antiemetic management contribute to the observed sex disparity in PONV.

### Heterogeneity and Influence Analyses

Due to the high heterogeneity (I^2^=93.1%) observed across the studies, both a Baujat plot and a leave-one-out sensitivity analysis were used to investigate whether any single study was contributing significantly to heterogeneity.

The Baujat plot (Figure 5) identified the study by Chae *et al*. as having both the highest contribution to heterogeneity and the highest influence on the overall result (14). However, leave-one-out sensitivity analysis (Figure 6) shows that the pooled effect remained stable across omissions of every study (OR range=2.31-2.49; overall 2.4). Omitting Chae *et al*. had a minimal effect on the pooled effect, resulting in OR=2.38 (95% CI [2.00-2.82]), and reduced heterogeneity from I^2^=93.1% to 89.2%.

**Figure 5.**
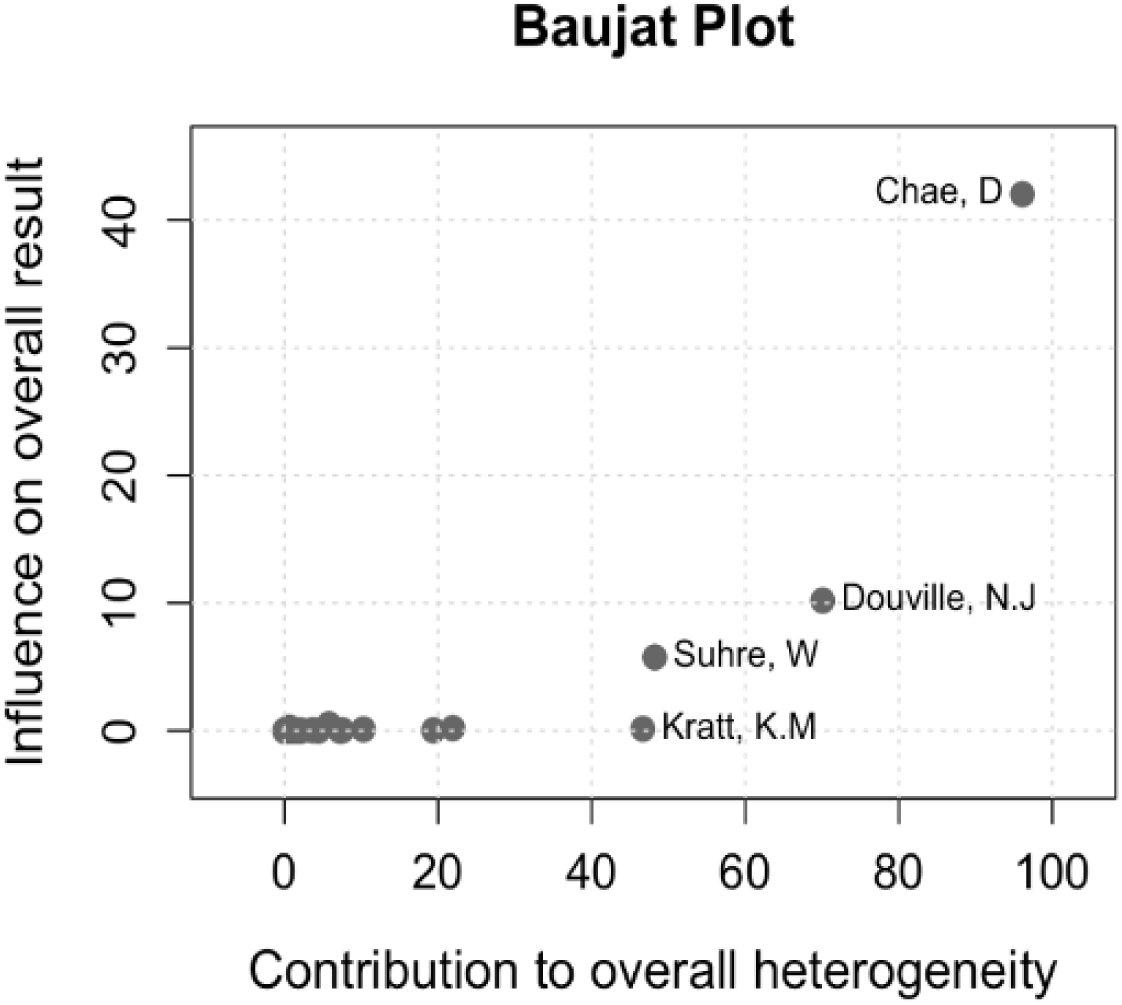
Baujat plot assessing study influence on heterogeneity and pooled effect. For legibility, studies with low contribution have not been individually labelled.

**Figure 6.**
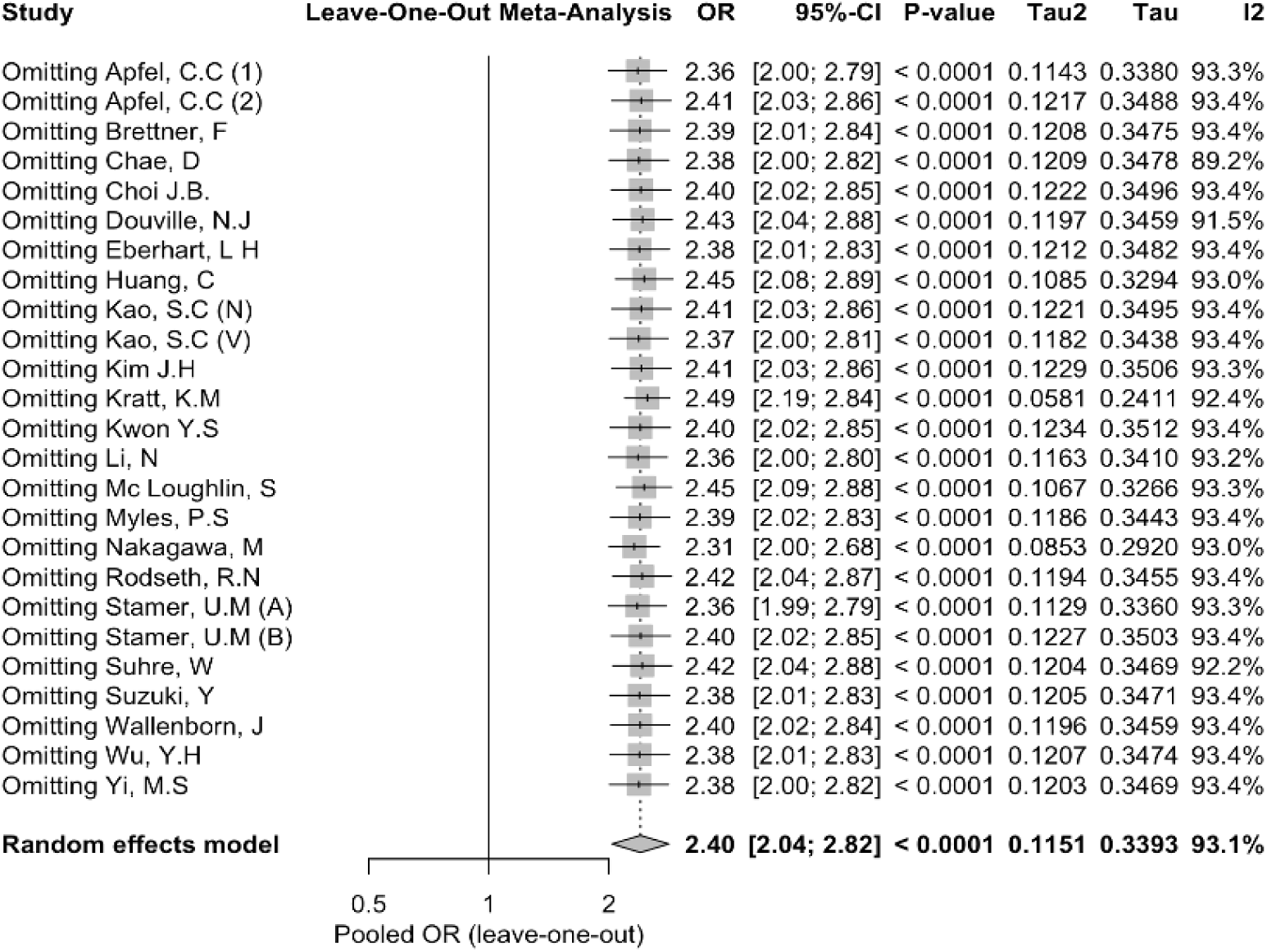
Leave-one-out sensitivity analysis. Influence of individual studies on the pooled OR for risk of PONV in females. Each line represents the pooled effect estimate after sequential omission of one study, demonstrating the stability of the overall result. Two studies by Apfel et al. are labelled (1) and (2). Kao et al. reported separate nausea and vomiting datasets, labelled (N) and (V), respectively. Stamer et al. included two independent cohorts, labelled (A) and (B).

The leave-one-out sensitivity analysis further showed that omitting Kratt *et al.* had the greatest upward shift (OR=2.49 (95% CI[2.19-2.84)], reflective of Kratt *et al*. as the only study that reported females having a lower incidence of PONV than males (OR=0.73 95% CI[0.52-1.03]) (11). Omission of Nakagawa *et al*. produced the greatest downward shift (OR=2.31 (95% CI[2.00-2.68]), which is consistent with this study showing the highest OR from its multivariate regression analysis (OR=7.26 95% CI[4.44-11.88]) (15).

### Risk of Bias Assessment

Most studies were rated as having high concerns for risk of bias. Overall, six studies (26.1%) were rated as having some concerns of bias, 11 studies (47.8%) were rated as having high levels of bias, and six studies (26.1%) were rated as having very high levels of bias. The domain that contributed most to increasing risk of bias was “bias in selection of participants into the study (or into the analysis)”. Risk of bias in other domains was generally rated as “low” or having “some concerns”, with very few ratings of “high”. There were concerns about bias in the measurement of PONV in most studies due to varying methods, including staff observations, the use of rescue antiemetics, and patient reporting of outcomes. Full domain-level and overall assessments for each study are presented in Figure C-1 (Appendix C).

## Discussion

This systematic review and meta-analysis found that females are at consistently more than double the risk of developing PONV than males. Despite high heterogeneity across studies, a remarkable consistent effect size was shown across studies including across sensitivity and subgroup analyses, suggesting that the variation observed in study findings is likely due to population differences and not random error. This was reflected across 22 out of the 23 studies included in the analysis, which encompassed a wide range of surgical settings, countries, and patient ethnicities. These findings underscore the need for further research to understand the mechanisms underlying this consistently increased risk of PONV in females, to inform targeted strategies, that may include more equitable use of PONV prophylaxis across sexes to address the demonstrable disparity in perioperative outcomes.

The findings of this systematic review support the current understanding of the female sex as one of the most significant non-modifiable, patient-specific risk factors of developing PONV (3,4). While female-exclusive surgeries, such as breast and gynaecological procedures, have been proposed to increase PONV risk, the sustained elevation in OR across all subgroups, including those excluding female-specific surgeries, suggests that this disparity cannot be solely attributed to surgical type alone. Instead, these findings may point toward underlying biological and physiological differences. Hormonal influences, particularly fluctuations in oestrogen and progesterone, have been widely proposed as potential contributing factors (16,17). Both oestrogen and progesterone can affect gut motility and sensitivity through direct activation of receptors; however, the exact link to PONV remains unclear (18,19). A systematic review by Eberhart *et al*. investigated the effect of the menstrual cycle on the incidence of PONV and found inconsistent results, suggesting that the menstrual cycle may not have an effect on the risk of developing PONV [(20)]. Furthermore, the absence of a similar sex-based disparity in paediatric and older populations supports a potential, but incomplete, hormonal contribution (21).

In addition to hormonal influences, other mechanisms such as sex-based differences in pharmacokinetics and pharmacodynamics of anaesthetic agents and perioperative pharmacotherapy are also hypothesised to contribute to this disparity (22). The higher ORs observed at 24 and 48 hours post-operation PONV end-points compared to the immediate postoperative period during PACU may potentially be related to differences in drug metabolism and response to anaesthetic agents (23). Females have been shown to exhibit increased sensitivity to opioids and slower induction but faster emergence to volatile anaesthetics, both of which are established risk factors for PONV, potentially increasing females’ susceptibility to PONV (24). Sex-based differences in the chemoreceptor trigger zone in emesis have also been proposed, although evidence remains limited (25). Taken together, the consistency of findings across surgical types, timepoints, and sensitivity analyses in this review supports a multifactorial explanation, involving an interplay of hormonal, pharmacological, and neurobiological factors. This complexity underscores the importance of recognising female sex as an independent and clinically meaningful risk factor, warranting targeted consideration in both risk stratification and prophylactic management strategies.

This review has several key strengths. Restricting inclusion to studies reporting adjusted estimates from multivariable analyses reduces confounding and strengthens internal validity. Large sample sizes (n ≥ 500) improve statistical power and precision. The use of random-effects modelling and sensitivity analyses enhances robustness, while inclusion of large observational studies supports real-world applicability.

The main limitations of this review include high heterogeneity across studies, mitigated by leave one study out and robust sensitivity analyses, as well as the generally increased risk of bias of included studies and potential publication and language bias due to restriction to English-language studies. A substantial proportion of bias arose from participant selection, with many studies limited to single centres, specific ethnic groups, or particular surgical settings. However, the overall diversity of included populations makes systematic distortion of pooled estimates less likely. There was considerable methodological and clinical heterogeneity, particularly in the measurement and timing of PONV, with varied and often subjective definitions and limited assessment of severity, hindering comparability across studies. Variation in covariates included in multivariable models may also limit comparability of adjusted estimates and introduce residual confounding. Further analyses by patient factors such as ethnicity and age were not possible due to limited reporting. Additionally, insufficient data were available to assess differences in PONV prophylaxis between sexes, as most studies did not stratify prophylaxis use, indicating an important area for future research.

This systematic review and meta-analysis showed that females are at significantly higher risk of developing PONV compared to males. Despite this well-established risk, there remains a poor understanding of the underlying mechanisms. Further work is now needed to tailor prophylactic and therapeutic strategies to this demonstrable disparity. The findings emphasise the need for sex-specific reporting, including on the use and effectiveness of antiemetics, alongside adherence to prophylaxis protocols. In conclusion, this systematic review highlights the need to address this sex-gap to ensure more equitable and effective management of PONV in clinical practice.

## Appendix A Search Strategy

### Search strategy

The databases searched for this review were Medline (OVID) and Embase. The search combined four key concepts using Boolean operators: postoperative nausea and vomiting (PONV), sex (female and male), and multivariable regression analysis. Searches were limited to English-language studies and excluded animal studies using database filters. Additional relevant studies were identified through manual screening of reference lists.

The specific search strategy is shown below.

”Postoperative Nausea and Vomiting” OR (post?surg* nausea or post?anesthe* nausea or post?op* emes?s or PONV or post?op nausea). mp. AND Female/ OR (wom* or female* or biologic* female* or AFAB or cisgender wom*).mp. AND Male/ OR (men or male* or biologic* male* or AMAB or cisgender m*).mp. AND exp Regression Analysis/ or exp Multivariate Analysis/ or ((multivariate or logistic or linear) adj3 (regression or analysis or model)).ti,ab.

## Appendix B Pooled Incidence of PONV

**Figure B-1.**
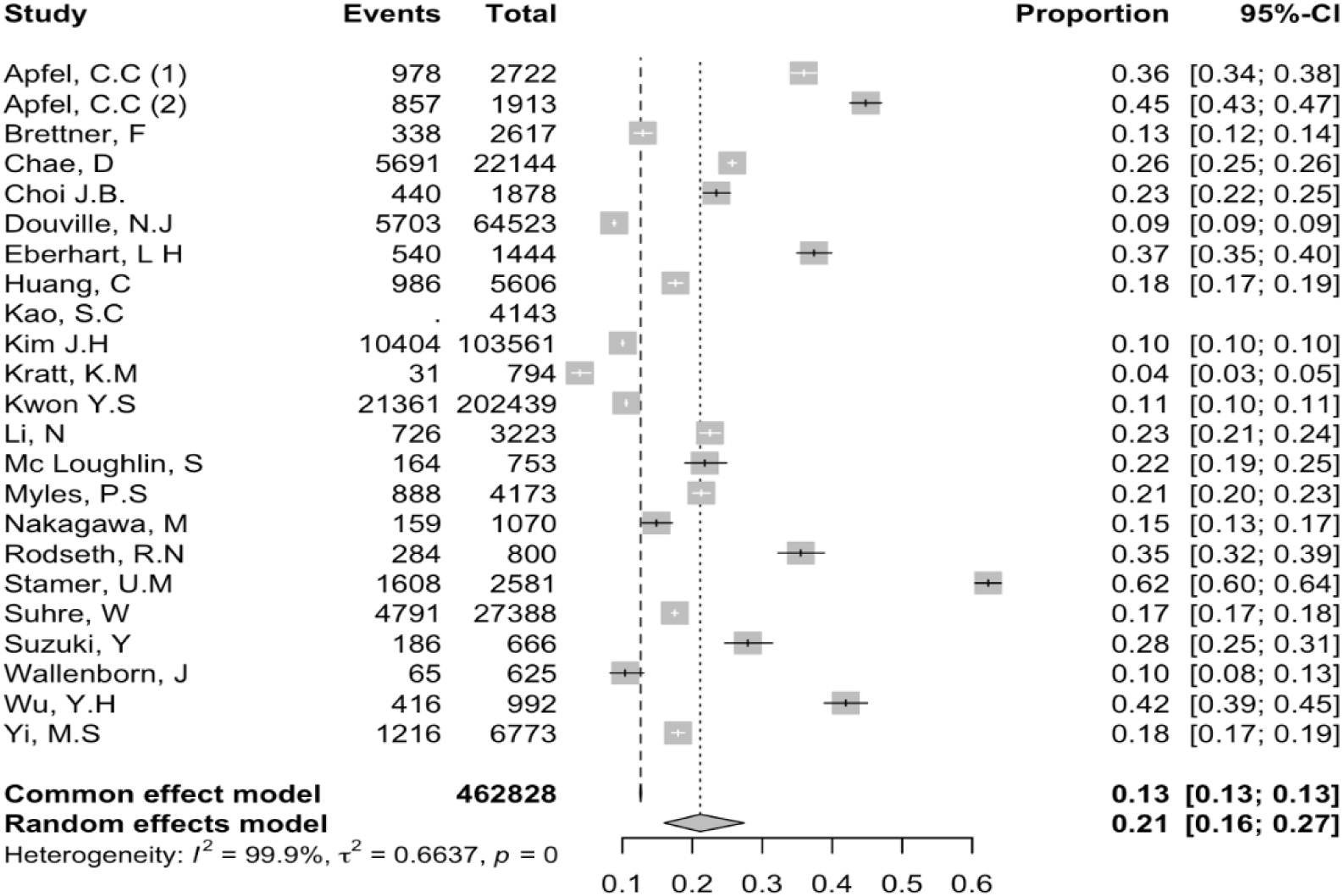
Pooled incidence of PONV across all included studies. Events = number of patients who developed PONV; Total = total study population. Two studies by Apfel et al. are labelled (1) and (2).

## Appendix C Risk of Bias (ROBINS-E) Assessment

**Figure C-1.**
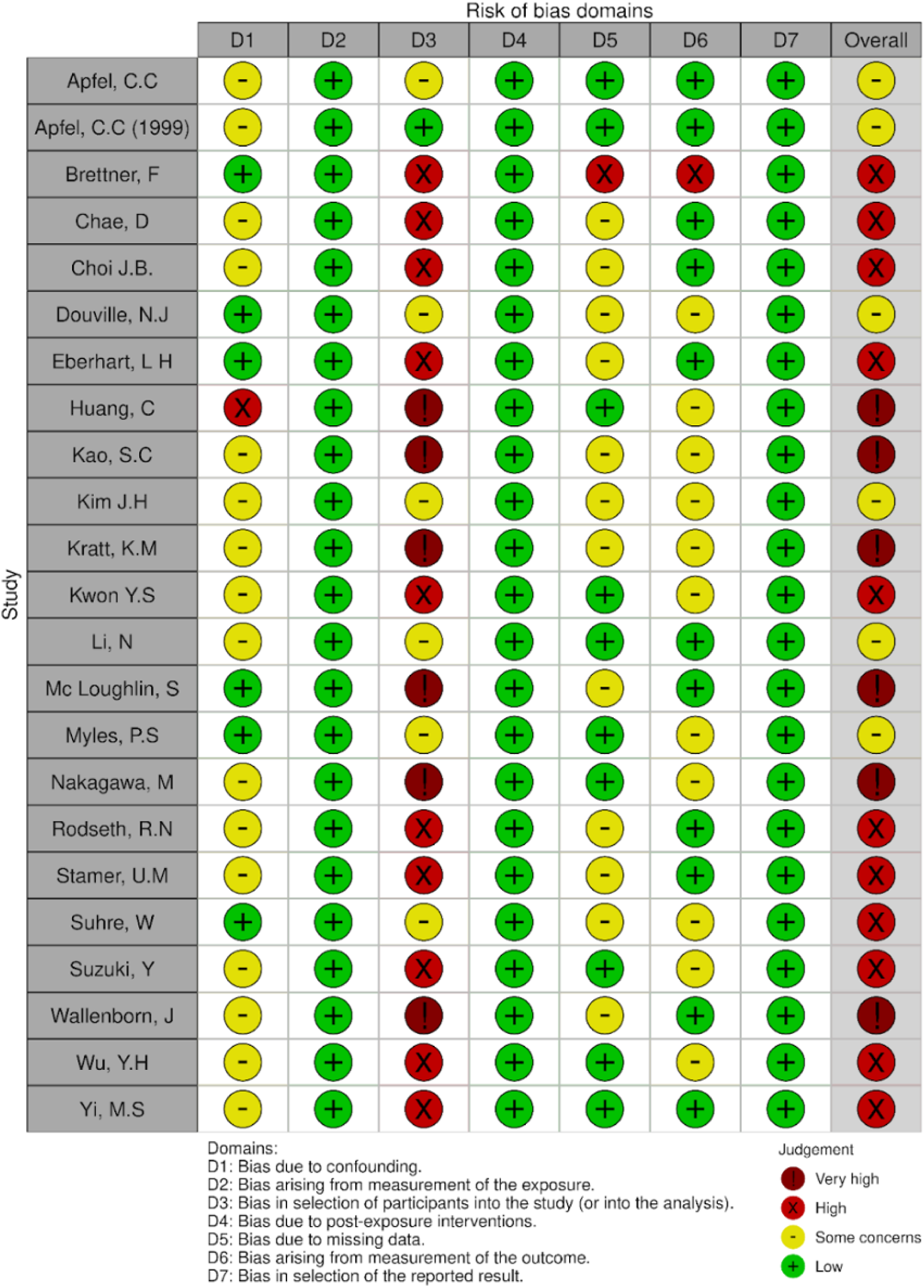
ROBINS-E Risk of Bias Assessment Outcomes.

## Data Availability

All data produced in the present study are available upon reasonable request to the authors

## Notes

**Funding:** This study was funded by the Health Research Council of New Zealand.

### Competing Interest Statement

GO is a co-founder and CEO of Alimetry Ltd. and has various grants and patents in the gastrointestinal field. ML and CV are affiliated with Alimetry Ltd.

### Author Declarations

This systematic review included publicly available published articles that analysed patient data.

